# Applied machine learning techniques for chronic disease treatment default prediction and its potential benefits for patient outcome: A case series study approach

**DOI:** 10.1101/2024.01.02.24300700

**Authors:** Michael Owusu-Adjei, Gaddafi Abdul-Salaam, Twum Frimpong, James Ben Hayfron-Acquah

## Abstract

In medical diagnosis context, consideration for missing diagnosis and false diagnosis of disease types are important clinical considerations for disease treatment decisions. Effect and impact of disease types especially on others forms the basis for critical clinical decisions. Impact and consequences varies across disease types especially for communicable and non-communicable diseases. Increasing use of predictive techniques owing to high use of connected internet of things devices in healthcare provides sufficient opportunity for potential benefit assessment of predictive modeling impact on disease treatment management. Effective and efficient management of non-communicable diseases such as hypertension is hampered in part by instances of multiple forms of its occurrence in patients leading to treatment management complications. Probing predictive modeling effect and implications for clinical decisions to enhance patient treatment outcome provides important evidence-based justifications for its use in healthcare systems. Effective predictive technique use is significantly dependent on areas of its application and the consequences of error for its use in context.

**Author summary:** The use of macro average score in class imbalance context is to treat all classes equally regardless of variations in class distributions. Clinical significance as identified in this research work includes the determination of effective and accurate predictive modeling techniques for real-world application context where class distribution variation is a characteristic feature. Identifying various sections of healthcare delivery process ensures effective application of predictive modeling techniques for the required impact on clinical decisions and its effect on patient outcome.

## 1.0 Introduction

Effect and impact are important clinical considerations for disease treatment decisions in healthcare. Online Cambridge dictionary defines impact as “the force or action of one object hitting another” or “a powerful effect that something, especially something new, has on a situation or a person”. The resulting influence of action or force is determined to be the effect of impact. Crucial to effective patient care is, evidence-based clinical decisions which includes fast, intuitive and deliberate analytical decision considerations within the context of disease type (communicable, non-communicable), severity, effect and impact on patient care and others [1]. Clinical adoption of modeling techniques is dependent on two most important factors; clinical usefulness and trustworthiness. Model technique role in clinical practice and expected outcome must therefore answer specific clinical questions with attendant patient benefits else its usefulness and trust for result reliability and dependability is questioned [2][3]. Effective application of predictive techniques in medical context consists of quality and accuracy of data given, assessment of effect on clinical decisions and impact on patient outcome taking into account disease type. The lack of performance assessment in clinical contexts is amplified in a related study [4] that examined machine learning tools in two areas; sepsis diagnosis and suicide prediction. Research on development of computerized clinical decision support systems have seen a tremendous growth in recent times but research assessment on specific learning techniques and use and potential benefit to clinicians and patients remains a challenge. These challenges as identified are mainly driven by concerns on accountability of usage in unfavorable clinical decision outcomes, risk of over-reliance due to limitations and errors and the problem of usability involving integration into existing clinical workflows [5][6]. Context-based predictive modeling assessment for potential benefit to clinical decisions on patient outcome in chronic disease treatment with comorbidity remains limited in research. This research work brings to the fore important clinical decision points for predictive modeling technique performance impact assessment on both clinical decisions and patient outcomes in chronic disease treatment default prediction.

### 1.1 Research objective

The key research objective is to evaluate the impact of predictive performance and its effect on clinical decisions in patient outcomes in chronic diseases such as hypertension with comorbidities’ for potential benefits. The outcome of this research study will help address impact and effect of predictive technique use in clinical decisions and patient outcome.

### 1.2 Research significance

The burden of chronic diseases and its negative impact on health management especially in the lower and middle income countries continue to attract attention from both policy makers and research studies. Limited number of healthcare professionals to adequately manage the increasing surge and its negative impact especially in developing countries where it’s devastation is affecting economies, increasing healthcare costs and patient morbidity resulting in increasing patient deaths. Trust and believe in predictive modeling technique use as a reliable alternative has the potential to help address human resource challenges. It is therefore significant to determine predictive modeling performance impact and effect on clinical decisions and patient treatment outcome that build confidence in its application particularly in healthcare systems.

### 1.3 Related research works

To help address potential benefits of applied predictive technique performance on clinical decisions and its effect on patient outcome, the use and impact on clinical decisions is examined in related applications in various research works particularly on communicable, non-communicable and other important considerations. The importance and use of predictive techniques in accelerating and enhancing physician tasks through effective automation processes has also enhanced and improved decision making in healthcare as emphasized in research studies [7][8] that examined machine learning applications in healthcare particularly public health. Furthermore, predictive algorithm use and its impact include significant effect on accurate predictions that positively impacts on clinical decisions and treatment outcome such as disease diagnosis and prognosis, personalized medicine, public Health surveillance and outbreak detection, health behavior analysis and intervention and healthcare resource utilization. Some of the benefits as determined in a related study show how data-driven insights and informed decision-making with machine learning use has revolutionized healthcare and public health decision support systems. In public health delivery, predictive technique use has been beneficial in spatial modeling, risk prediction, misinformation control, public health surveillance, disease forecasting, pandemic/ epidemic modeling and health diagnosis [9]. Other application areas for predictive technique use as identified include precision medicine, diagnosis and treatment recommendations, patient engagement and adherence and administrative activities [10]. In their article on Artificial intelligence, machine learning and health systems, the potential effect of machine learning to be the catalyst for healthcare systems improvement in efficiency, effectiveness and outcome is emphasized [11]. Another scoping review of clinical decision making in the emergency department for paediatric patients [12] include among its focus models for clinical decision support. A randomized trial of 20, 563 patients admitted to a hospital were assessed to determine the effectiveness of computerized clinical decision support system. This provided evidence-based appraisal for actionable, patient-specific recommendations at the point of care by healthcare professionals [5]. The need for model accuracy improvement within an acceptable time interval for early detection of diseases such as cancer, diabetes, chronic kidney diseases etc is underscored in a research study which compared performance of different machine learning techniques and approaches for clinical decisions [13]. Identified resource areas for predictive application decisions in healthcare were in patient care, disease treatment, resource allocation and utilization. Results obtained indicate that using neural network-based deep learning techniques in computational biology domains produced high prediction accuracy with reliability, thus making computational biology and biomedicine-based decisions dependent on such predictive modeling techniques. Similar use of machine learning techniques for the prediction of Diabetes Mellitus Disease with feature augmentation and oversampling techniques [14] showed high prediction accuracy performance in two distinct datasets for classifiers such as random forest (RF), light gradient boosting (LGB) and gradient boosting (GB). Prediction accuracy scores obtained were 98.99% for LGB, 96.6% for RF and 97.64% for GB. Conclusions drawn was a proposal for further improvements in prediction accuracy with advanced methodologies such as transformer-based learning technique. Applied machine learning techniques for cervical pain assessment on patients affected by whiplash disease using techniques such as logistic regression, support vector machines, k-nearest neighbors, gradient boosting, decision trees, random forest, and neural network algorithms on 302 dataset examples [15] produced prediction accuracy, precision and recall values above 90%. Predictive performance benefit and clinical impact and patient effect as identified include presence of pain in patients for clinical decisions that can improve treatment efficacy to help address resource utilization. Comparative study to predict measles with 1,797 suspected cases used six machine learning techniques to determine 78 positive cases for measles and 1,696 were identified as negative cases creating a class distribution imbalance. Results obtained showed superior random forest classifier performance in specificity 96%, sensitivity 88%, receiver operating characteristic curve score 92% and total prediction accuracy score 92% [16] than the other modeling techniques (generalized linear model, decision tree, naïve bayes, support vector machines, artificial neural network). Applied learning technique assessment study for the detection and management of infectious diseases caused by fatal or life-threatening causative agents capable of infecting both animals and humans provided a comprehensive review of machine learning application use in pathogen detection, public health surveillance, host-parasite interaction, drug discovery, omics and vaccine discovery. Emphasis on the use of evaluation metrics (precision and recall) in classification tasks [17] as important performance measure is made in respect of modeling techniques such as support vector machine, random forest and neural networks. Clinical impact is assessed to be the inclusion and emphasis on precision and recall as important evaluation metrics to identify parasitic cells as infected or not infected (positive or negative). Impact of multivariate algorithm use [18] to accurately select breast cancer patients without residual cancer after administering neo-adjuvant therapy is identified in a related study that identified breast cancer patients with pathologic complete response. Using randomized dataset examples of 457 women, partitioned into training and test set enrolled in three trials with stage 1-3 breast cancer, False-negative rate (FNR) estimate of 1.2% for logistic regression with elastic net penalty, extreme gradient boosting, support vector machines and neural network was achieved. Estimating the incidence of diabetes mellitus in a public health surveillance based on the number of reimbursements over a 2 year period [19] with four predictive techniques; Linear discriminant analysis (LDA), Logistic regression, (LR), Flexible discriminant analysis (FDA) and Decision tree for 44,659 participants showed predicted performance score of sensitivity 62%, specificity 67% and accuracy score of 67% for LDA, making it the highest performing technique of choice. Effective modeling techniques in healthcare as identified [20] in a comparative analysis of modeling technique study include random forest classifier. Context-based predictive modeling assessment especially in real-world business applications should therefore explore specific areas in the clinical decision process for relevancy to determine potential benefits and the consequences of error as reasonable justification for its use.

## 2.0 Methods and Materials

In this study, a case series approach was adopted using characteristics identified from 5,333 electronic health records of patients undergoing treatment for hypertension with comorbidity. Used variables were extracted from patient attendance interactions as recorded by attending clinicians with expressed permission request and approval referenced DCS/S.1/Vol.1 and dated 30^th^ march, 2022 from the main district healthcare facility in Kwahu South, Ghana. The reason for selecting this healthcare facility is informed by its long standing management of cardiovascular diseases including hypertension, its status as a referral point and remote location serving several communities with diverse population characteristics including education, occupation, social behavior, health needs and many others. The uniqueness of its location within a mountainous geographical location of Ghana with variable weather conditions characteristically different from other parts of Ghana makes it the ideal location for insights into such phenomena. Among the sampled patients, 4,312 constituting 80.86% were female and 1,021 constituting 19.14% were male. Patients with hypertension but no comorbidity was 4,192 making up 78.60% and 1,141 were identified patients with both hypertension and comorbidity constituting 21.40%. In this study, gender feature was separated into male and female, incidence of comorbidity among patients was also separated into patients with only hypertension without comorbidity and patients with both disease conditions (hypertension with comorbidity). Selection criteria for inclusion and exclusion are defined below.

### 2.1 inclusion criteria

1. Criteria for inclusion included patients diagnosed of hypertension with and without comorbidity.
2. No exclusivity rule was applied in respect of gender preferences, age, social status, occupational preferences etc.
3. Patients who had been diagnosed from 6 months and above were considered.
4. Patients enrolled on National health insurance and other private personal insurance including those not covered by any insurance policy were considered.
5. No consideration for the number of comorbidity instances in a single patient.
6. Patients diagnosed and profiled as defaulters and non-defaulters were included.

### 2.2 exclusion criteria

1. Transfer-in patients regardless of length of stay was excluded.
2. Pregnant patients with any of these disease conditions were excluded
3. Disabled patients with challenges in movement were excluded
4. Diagnosed patients with mental health disorders were excluded

#### Ethical approval and Consent

Real-world clinical dataset obtained from the main district hospital in Kwahu South district of Ghana with approval notice reference number DCS/S.1/VOL.1 dated on 30^th^ march, 2022.

#### 2.0.1 Methodology

Determination of specific areas in the decision process for potential benefits in the use of predictive techniques remains the main research objective in this paper. Determining model performance impact in such areas will provide important insight for clinical decision support systems for effective healthcare management that improves patient outcome. Ten (10) known modeling techniques namely; logistic regression, support vector machine, random forest classifier, gradient boosting classifier, multi-layer perceptron, extra trees classifier, decision tree, bagging classifier, kneighbors classifier and linear discriminant analysis classifier are employed to estimate predictive performance and how it impacts on clinical decision and its effect on patient outcome. Brief but less technical description of the predictive techniques used are presented in the following sub-sections.

#### 2.0.1.1 Logistic regression

Predominantly about 70% of problems in data science and data mining are classification problems for which classification techniques have become essential part of machine learning applications [21]. Logistic regression use in binary classifications is evident in spam detection, disease diagnosis and predictions such as diabetes, chronic kidney, cancer predictions, tumor classifications into malignant and benign, customer purchasing behavior predictions etc. its use is particularly important in establishing relationships between dependent and independent variables in two-binary classifications.

#### 2.0.1.2 Support vector machines

Its ability to handle non-linear input space and separate data points using hyperplane that finds optimal data points to classify new data points. Reference to support vector machine as a discriminative classifier [22] makes it an ideal technique for intrusion detection, face detection, email classifications, etc. it is useful in both classification and regression problem domains.

#### 2.0.1.2 Random forest classifier

Random forest classifier is a supervised learning technique used in classification, regression and other tasks with decision trees. Decision trees are created from randomly selected subset of training sets. Collection of votes from different decision trees determines final prediction outcome [23].

#### 2.0.1.3 Gradient boosting classifier

Structured predictive modeling problems such as classification and regression find it useful on tabular data modeling [24]. It is an ensemble algorithm that minimizes error gradients with fitted boosted decision trees. It is also known as gradient tree boosting, stochastic gradient boosting and gradient boosting machines and abbreviated as GBM.

#### 2.0.1.4 Multi-layer perceptron

Multi-layer perceptron or MLP is a type of neural network algorithm used mostly in complex classification tasks. Its capability lies in its ability to learn non-linear relationships between inputs and outputs [25]. MLP has an input layer to receive inputs, hidden layer to perform computations on inputs received and an output layer to generate final results for display from the model.

#### 2.0.1.5 Extra trees classifier

Classification and regression task algorithm that works by selecting a subset of features randomly for training by a decision tree. The tree is subsequently pruned [26] to contain features important for predicting the final outcome. It is also known as Extreme randomized tree.

#### 2.0.1.6 Decision trees

Supervised learning models useful in both classification and regression problems. The work in a decision tree is representative of a flowchart in which each node represents a decision point that splits into two leaf nodes representing an outcome of a decision. Each of these decisions can also turn into decision nodes [27]. The decision making process in decision trees are easy to understand and interpret. It works by splitting data into series of binary decisions for traversal.

#### 2.0.1.7 Bagging classifier

An ensemble algorithm that works by dividing training sets into subsets for processing by different techniques to improve performance. Combined outputs from the different techniques form the basis [28] for performance determination. Bagging is useful in classification and regression problem domains.

#### 2.0.1.8 Kneighbors classifier

Supervised machine learning algorithm predominantly used in both classification and regression tasks. Predictions are made by calculating the distance between the two data points (training data and test data) for the assumption of characteristics that is similar between them [29]. Characteristics learned from the training data points allow for the identification and allocation of new data points.

#### 2.0.1.8 Linear discriminant analysis classifier

Useful supervised learning technique for statistics and classification. It works by finding linear combination features in a dataset class that best discriminates or separates them. It is particularly useful in finding informative features of importance in classification predictions. It is widely used in fields such as pattern recognitions, bioinformatics and image processing etc [30].

## Discussion

Statistical distribution of gender contributions to the incidence of hypertension with comorbidity is illustrated in **Table 1** which shows disease occurrence among age groups and gender including incidence of combidity. Among the female population, incidence of hypertension only is estimated at 3,435 and hypertension with comorbidity amounted to 877 within different age brackets. Estimated age range for only hypertension and hypertension with comorbidity is between 21 years to 109 years and 18 years to 111 years. Estimated age range for male population regarding incidence of hypertension only and hypertension with comorbidity was between 25-105 years and 23-100 years respectively. These descriptions show that incidence of hypertension and hypertension with comorbidity occur in female populations earlier than males and this is supported by research studies [31][32] that examined gender differences in the prevalence of hypertension across various age groups and potential association of gender-specific factors for hypertension. A comprehensive cardio-metabolic risk assessment of gender found females to be at high risk than males and among gender specific factors, age (younger age) at childbirth was found to be among the high risk factors for women. This study adds that the incidence of hypertension and hypertension with comorbidity occur in women 4-5 years earlier than men as shown in **Table 1**. Predictive model performance and its impact on clinical decisions and effect on patient outcome is accomplished in two different ways; **Table 2** show macro average model prediction precision, recall, f1-score and accuracy performance for the ten algorithms used. The importance and impact of each metric used is determined in context. As an example, it is better to misclassify a genuine email message as spam than to incorrectly misclassify a malicious spam message as genuine because of impact and effect. In medical contexts, misclassifying a compliant patient as a defaulter may have little impact and effect on clinical decisions and treatment outcome than misclassifying a default patient and this is significantly due to impact and effect on non-adherence. It is observed in **Table 2** that even though LDA achieves the highest area under the curve score (auc_roc) of 0.90 or 90%, its precision is hampered by attaining a score of 0.68 or 68%. However, logistic regression in this instance achieves the biggest precision score of 0.87 or 87% and an auc_roc score of 0.89 or 89%. However, macro average recall and f1-score as recorded show that random forest, extra trees, linear discriminant analysis and gradientboosting classifiers obtained the largest macro average recall score of 0.62 or 62% respectively as against 0.59 or 59% recorded for logistic regression and decision tree. The use of macro average scores in class imbalance context is to treat all classes equally regardless of variations in class distributions. Clinical significance includes a determination of effective and accurate predictive modeling techniques for real-world application context where class distribution variation is a characteristic feature. Identifying various sections of healthcare delivery process ensures effective application of predictive modeling techniques for the required impact on clinical decisions and its effect on patient outcome. Characteristic of real-world applications, collected dataset as displayed in **Fig 1** and **Fig 2** shows output class distribution imbalance between defaulted patients and non-defaulters. Estimated patient defaulters and non-defaulters amounted to 1.52% and 98.42% of the sampled population. In **Fig 3** important decision sections that serve as points for data collection in a typical healthcare system is provided and highlighted in red for emphasis. Sections highlighted provide useful context for predictive modeling application. Predictive modeling contribution for impact and effect in each of these sections towards clinical decisions and outcome remains varied therefore predictive research in any of these or combination of sections can provide important insight into its potential benefit for clinical decisions that enhances patient outcome as each of these sections show inter-dependency. In its estimation of nursing processes to serve as guide for nursing practice, subjective and objective assessment of critical thinking and data collection is determined as an important first step to achieving patient-centered care. Subjective assessment include verbal statements taking from patients and healthcare givers and objective assessment involves measurable metrics such as vital signs [33]. Clinical significance of nursing process utilization will ensure that the risk of missing out on hidden but life-threatening disease condition is reduced. Predictive technique use capable of learning patterns in clinical datasets offer important assistance to clinical decision support systems for patient management that improves treatment outcome. An outcome that include accurate classification and prediction of disease conditions, associated risk factors, identification of hidden patterns in different segments of large clinical datasets for effective prognosis of disease conditions. Model performance with receiver operating characteristic curve (roc_auc) for the ten algorithms is displayed in **Fig 4** that also show performance scores achieved by the individual techniques. Easy visualization of model performance as shown by the roc_auc score curve is displayed in **Fig 5**

**Fig 1.**
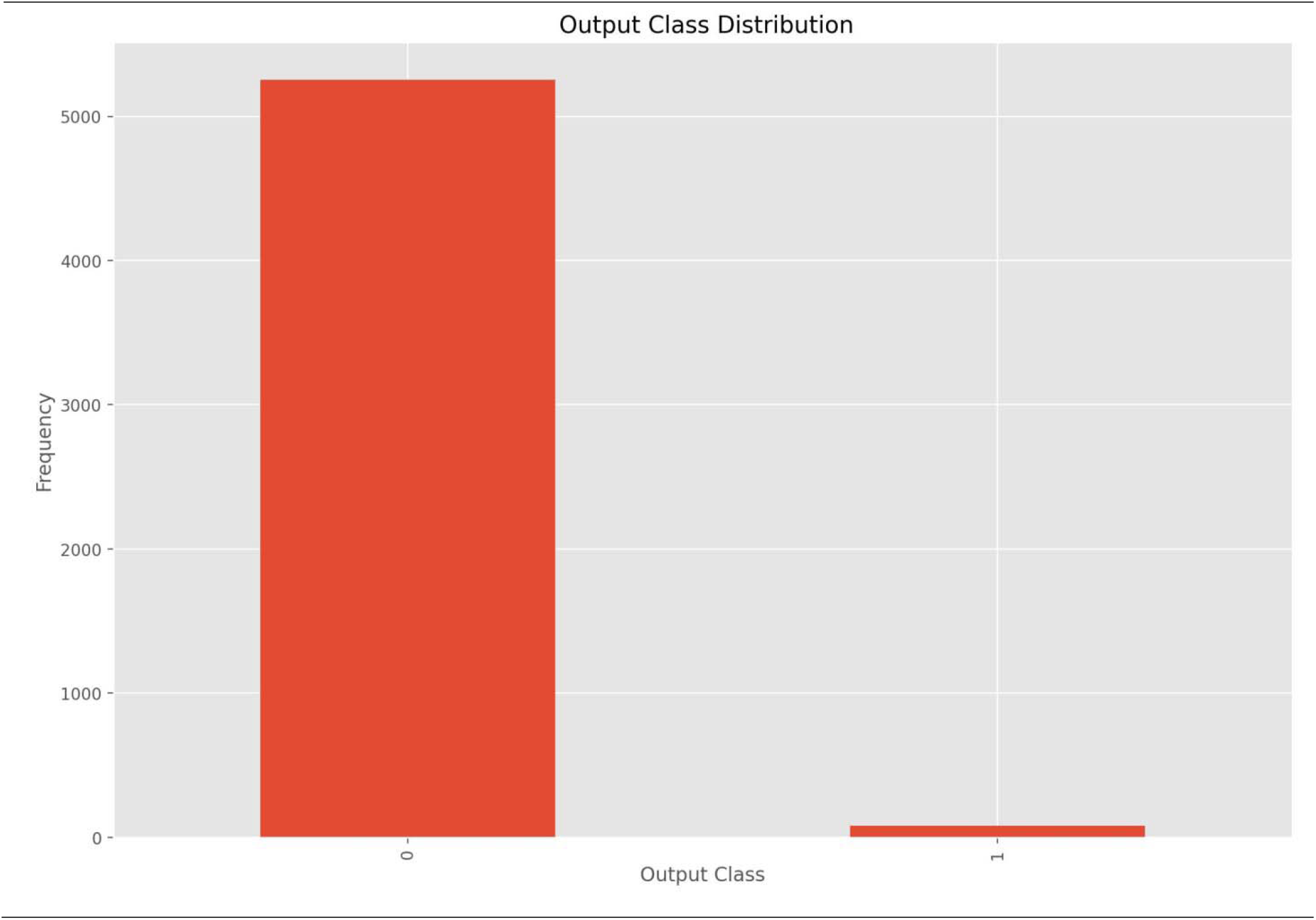
Output class distribution diagram. Caption: Output class distribution. This figure shows frequency distribution between defaulted and non-defaulted patients in the collected dataset.

**Fig 2.**
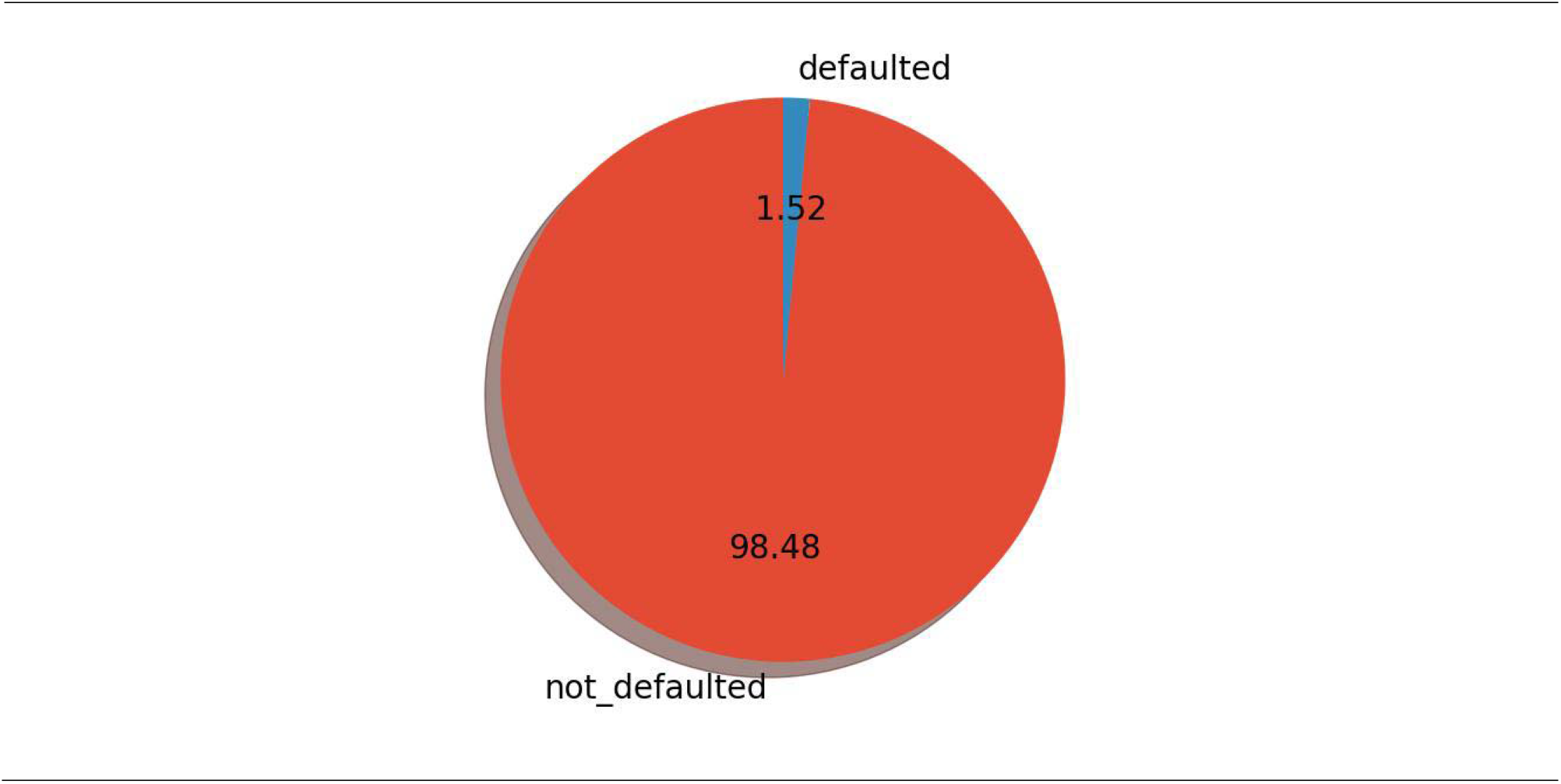
Distribution share of output class in percentage. Caption: Pie chart display of output class distribution. Distribution of percentage share in the output or target class is displayed. Percentage share of defaulters as estimated is 1.52% and non-defaulters is 98.48%.

**Fig 3.**
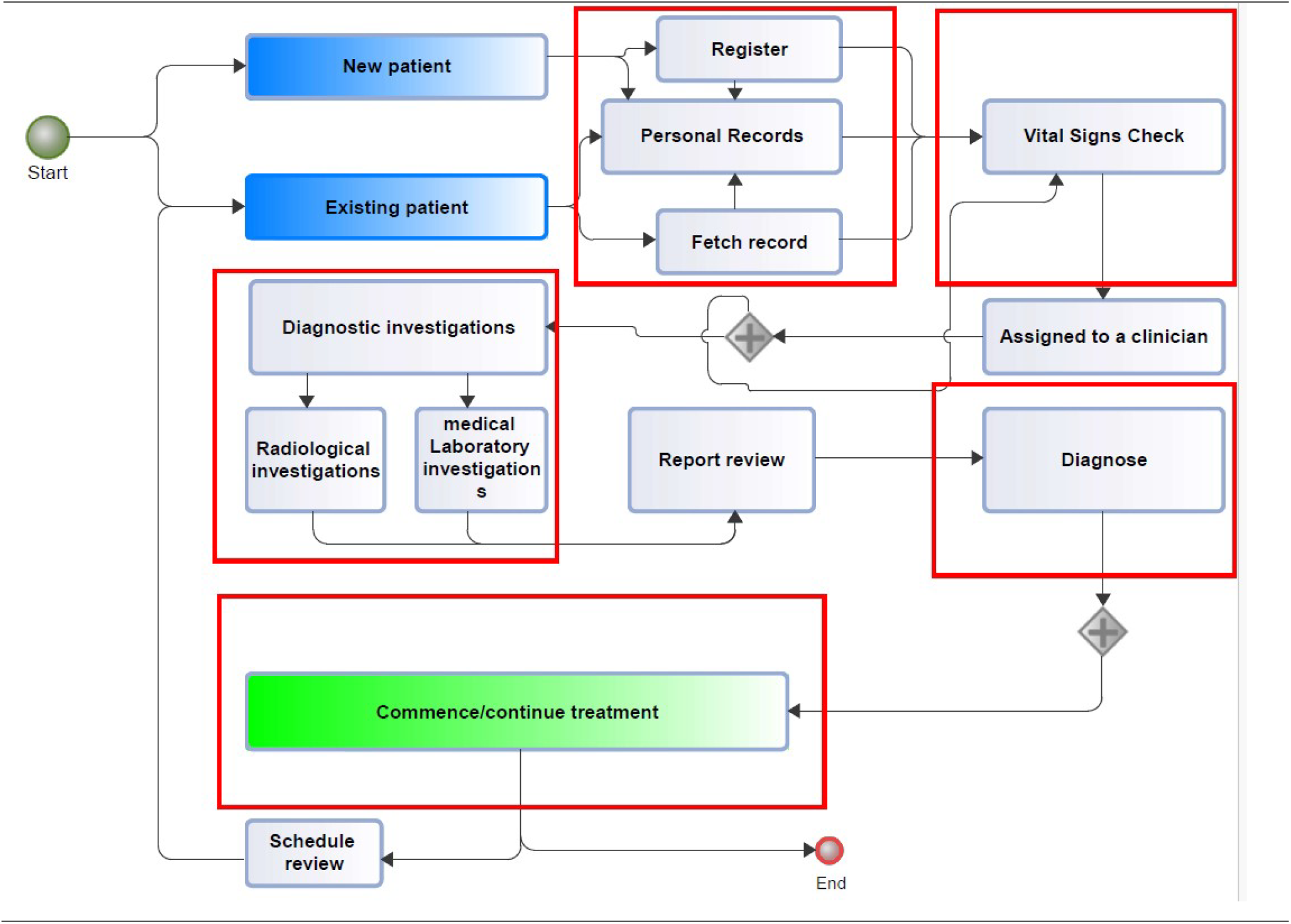
Clinical decision categories. Caption: Simplified clinical decision sections in a typical healthcare delivery system. Identified clinical decision sections relevant for data collection and applied modeling is marked and emphasized.

**Fig 4.**
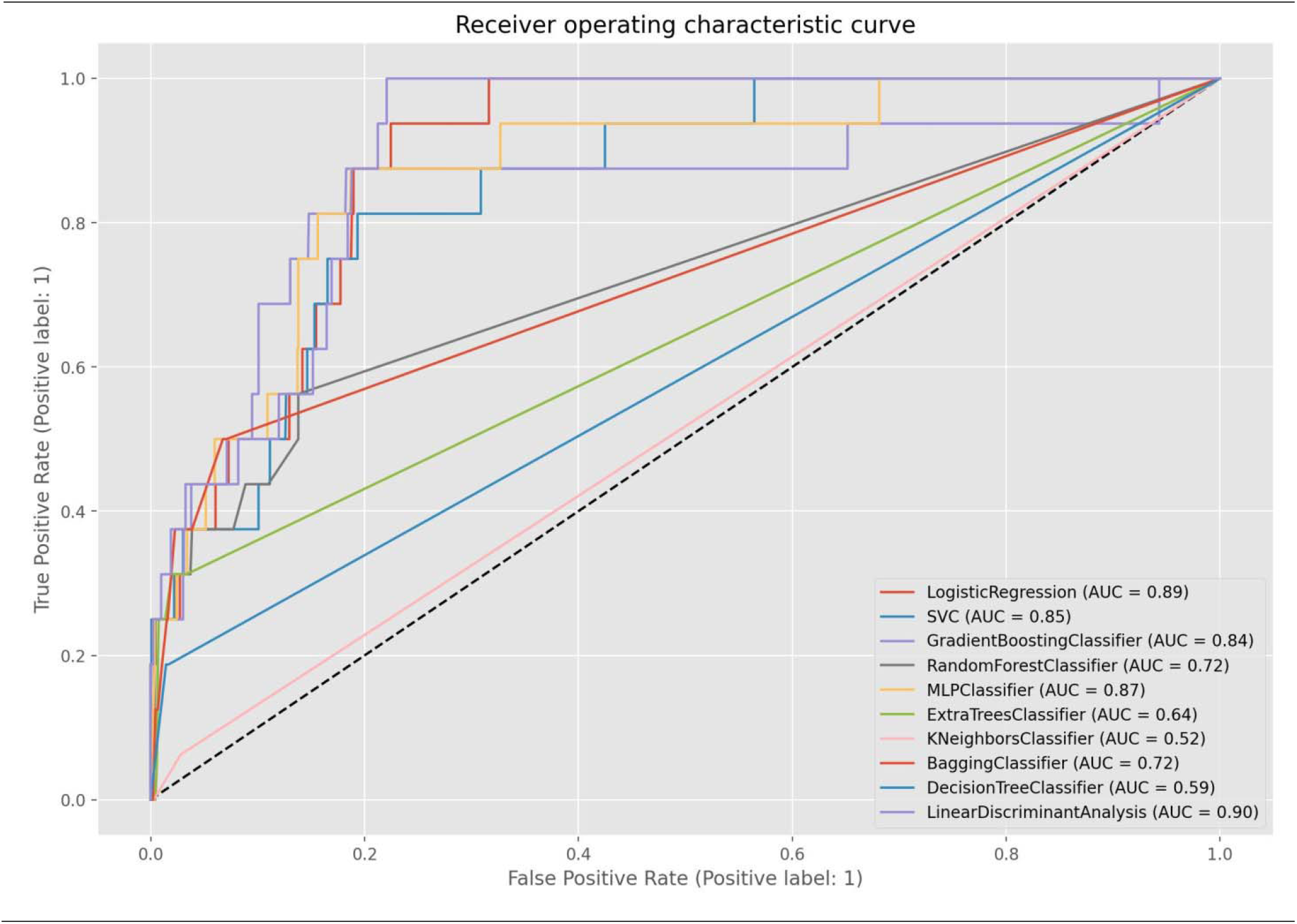
roc_auc score curve. Caption: Receiver operating characteristic curve. Evaluated model performance on predicted false positives and false negatives is determined by the use of roc_auc score curve.

**Fig 5.**
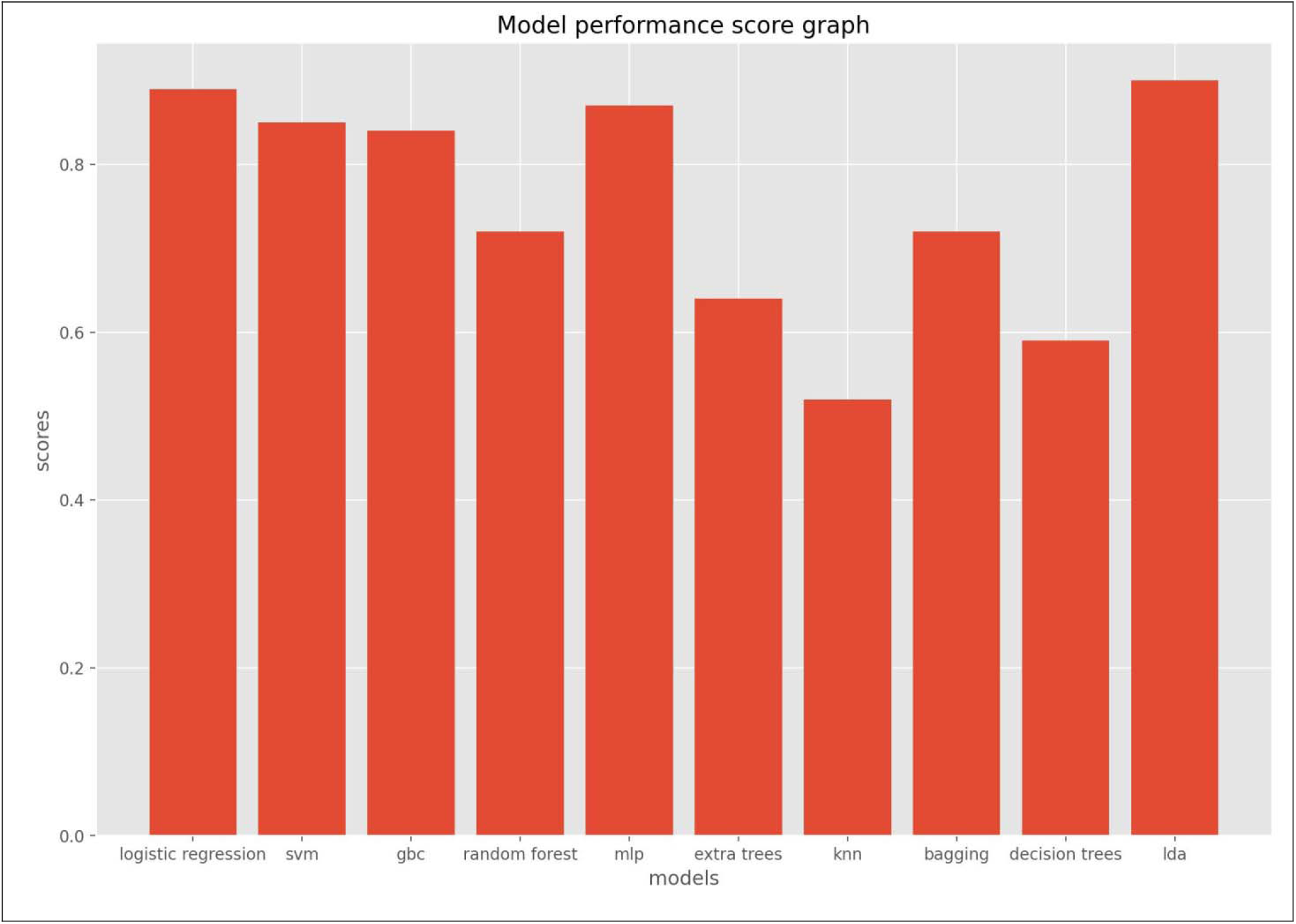
Individual Model performance display. Caption: Model performance score graph. This graph provides easy visualization to model performance scores as shown in the roc_auc score curve.

**Table 1.** Statistical distribution of gender contribution.

| Gender | Age range(years)<br>Hypertension only | Hypertension only | Age range (years)<br>hypertension with<br>comorbidity | Hypertension with<br>comorbidity |
| --- | --- | --- | --- | --- |
| Female | 21-109 | 3435 | 18-111 | 877 |
| Male | 25-105 | 757 | 23-100 | 264 |

**Table 2.**
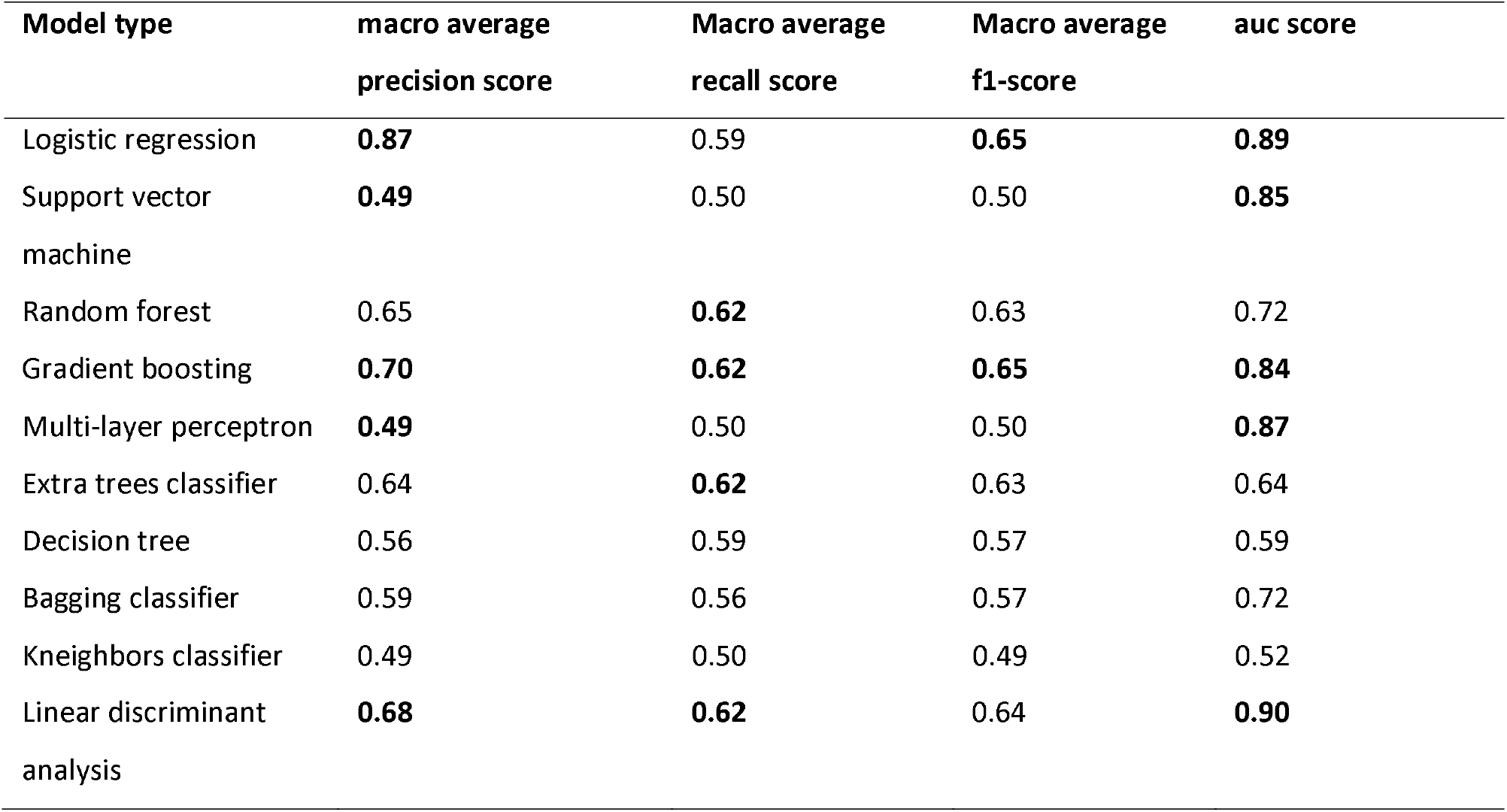
Model performance scores.

| Model type | macro average<br>precision score | Macro average<br>recall score | Macro average<br>f1-score | auc score |
| --- | --- | --- | --- | --- |
| Logistic regression | **0.87** | 0.59 | **0.65** | **0.89** |
| Support vector<br>machine | **0.49** | 0.50 | 0.50 | **0.85** |
| Random forest | 0.65 | **0.62** | 0.63 | 0.72 |
| Gradient boosting | **0.70** | **0.62** | **0.65** | **0.84** |
| Multi-layer perceptron | **0.49** | 0.50 | 0.50 | **0.87** |
| Extra trees classifier | 0.64 | **0.62** | 0.63 | 0.64 |
| Decision tree | 0.56 | 0.59 | 0.57 | 0.59 |
| Bagging classifier | 0.59 | 0.56 | 0.57 | 0.72 |
| Kneighbors classifier | 0.49 | 0.50 | 0.49 | 0.52 |
| Linear discriminant<br>analysis | **0.68** | **0.62** | 0.64 | **0.90** |

### Research study limitations

Any assumption made in this study is based on collected data available. Enhanced predictive performance especially for deep neural networks is dependent on large dataset volumes and this research dataset volume of 5,333 is considered minimal to deep neural network contexts for effective performance evaluation hence the use of traditionally tested algorithms. The inclusion of multi-layer perceptron classifier is for comparative assessment purpose. Identified class imbalance distribution of the output class is not addressed with any optimization techniques as its occurrence is characteristic of real-world applications.

### Research application and significance

The identification of decision sections offer predictive modeling research significant data collection points of interest for the creation of healthcare decision support systems. Prediction outcome with macro average precision, recall and f1-score determines how much trust to put into a prediction outcome in context which may define the impact of error on the spread of diseases for communicable and non-communicable diseases. Models with low macro average precision, recall and f1-score as recorded mean that the prediction of a default or non-default hypertensive patient cannot be trusted in such scenarios. This could lead to a situation where focus on default patients for necessary intervention is impaired impacting negatively on clinical decisions with attendant negative effect on patient outcome.

## Conclusion

The identification of significant decision sections that serve as important data collection points for impactful decision support in healthcare settings, determination of predictive technique impact in the use of relevant performance metrics within the context of real-world applications characteristic of output class distribution imbalance, exploratory analysis which show gender disparity in the incidence of hypertension and hypertension with comorbidity among different age groups demonstrates potential benefits for applied predictive modeling techniques on clinical decision impact and its effect on patient outcome. Future research will focus on collection and effective use of deep learning techniques for the diagnosis and interpretation of x-ray images as decision support system to address human resource challenges in the recruitment of qualified specialist radiologists in Ghana.

## Supporting information

Request and approval letter

## Data Availability

Research dataset used is available upon reasonable request

## Acknowledgements

Support and cooperation of both management and staff of Kwahu Government Hospital is acknowledged.

## Author contributions

**Conceptualization:** Owusu-Adjei Michael.

**Formal analysis:** Owusu-Adjei Michael.

**Methodology:** Gaddafi Abdul-Salaam.

**Project administration:** Owusu-Adjei Michael.

**Supervision:** James Ben Hayfron-Acquah

**Writing – original draft:** Owusu-Adjei Michael.

**Writing–review & editing:** Twum Frimpong.

## References

[1] T. J. Stein, “Critical thinking in clinical practice,” Child. Youth Serv. Rev., vol. 13, no. 5–6, pp. 441–443, 1991, doi: 10.1016/0190-7409(91)90032-d.

[2] S. Vollmer et al., “Machine learning and artificial intelligence research for patient benefit: 20 critical questions on transparency, replicability, ethics, and effectiveness,” BMJ, vol. 368, pp. 1–12, 2020, doi: 10.1136/bmj.l6927.

[3] S. A. Alowais et al., “Revolutionizing healthcare: the role of artificial intelligence in clinical practice,” BMC Med. Educ., vol. 23, no. 1, pp. 1–15, 2023, doi: 10.1186/s12909-023-04698-z.

[4] R. Kang, E. M. Rantanen, and E. A. Youngstrom, “Machine Learning in Healthcare: Two Case Studies,” Proc. Hum. Factors Ergon. Soc. Annu. Meet., vol. 66, no. 1, pp. 774–778, 2022, doi: 10.1177/1071181322661518.

[5] L. Moja et al., “Effectiveness of a Hospital-Based Computerized Decision Support System on Clinician Recommendations and Patient Outcomes: A Randomized Clinical Trial,” JAMA Netw. Open, vol. 2, no. 12, pp. 1–16, 2019, doi: 10.1001/jamanetworkopen.2019.17094.

[6] A. Choudhury and C. Allen, “Artificially Intelligent? Machine Learning in Healthcare and Why It May Not Be As Advanced As You Think,” Patient Saf., vol. 5, no. 2, pp. 76–83, 2023, doi: 10.33940/001c.77632.

[7] P. M. Rodrigues, J. P. Madeiro, and J. A. L. Marques, “Enhancing Health and Public Health through Machine Learning: Decision Support for Smarter Choices,” Bioengineering, vol. 10, no. 7, pp. 1–5, 2023, doi: 10.3390/bioengineering10070792.

[8] A. Zhang, L. Xing, J. Zou, and J. C. Wu, “Shifting machine learning for healthcare from development to deployment and from models to data,” Nat. Biomed. Eng., vol. 6, no. 12, pp. 1330–1345, 2022, doi: 10.1038/s41551-022-00898-y.

[9] D. B. Olawade, O. J. Wada, A. C. David-Olawade, E. Kunonga, O. Abaire, and J. Ling, “Using artificial intelligence to improve public health: a narrative review,” Front. Public Heal., vol. 11, no. October, pp. 1–9, 2023, doi: 10.3389/fpubh.2023.1196397.

[10] Z. Wen and H. Huang, “The potential for artificial intelligence in healthcare,” J. Commer. Biotechnol., vol. 27, no. 4, pp. 217–224, 2022, doi: 10.5912/jcb1327.

[11] T. Panch, P. Szolovits, and R. Atun, “Artificial intelligence, machine learning and health systems,” J. Glob. Health, vol. 8, no. 2, pp. 1–8, 2018, doi: 10.7189/jogh.08.020303.

[12] F. Leonard, D. O’Sullivan, J. Gilligan, N. O’Shea, and M. J. Barrett, “Supporting clinical decision making in the emergency department for paediatric patients using machine learning: A scoping review protocol,” PLoS One, vol. 18, no. 11, p. e0294231, 2023, doi: 10.1371/journal.pone.0294231.

[13] S. Mudalige, D. Alexis, and C. Jayatilake, “Involvement of Machine Learning Tools in Healthcare,” vol. 2021, 2021.

[14] B. Shamreen Ahamed, M. S. Arya, and A. O. Nancy, “Diabetes Mellitus Disease Prediction Using Machine Learning Classifiers and Techniques Using the Concept of Data Augmentation and Sampling,” Lect. Notes Networks Syst., vol. 516, pp. 401–413, 2023, doi: 10.1007/978-981-19-5221-0_40.

[15] J. de la Torre, J. Marin, S. Ilarri, and J. J. Marin, “Applying machine learning for healthcare: A case study on cervical pain assessment with motion capture,” Appl. Sci., vol. 10, no. 17, 2020, doi: 10.3390/app10175942.

[16] R. Gyebi et al., “Prediction of measles patients using machine learning classifiers: a comparative study,” Bull. Natl. Res. Cent., vol. 47, no. 1, 2023, doi: 10.1186/s42269-023-01079-w.

[17] R. S. Hu, A. E. L. Hesham, and Q. Zou, “Machine Learning and Its Applications for Protozoal Pathogens and Protozoal Infectious Diseases,” Front. Cell. Infect. Microbiol., vol. 12, no. April, pp. 1–17, 2022, doi: 10.3389/fcimb.2022.882995.

[18] A. Pfob et al., “Identification of breast cancer patients with pathologic complete response in the breast after neoadjuvant systemic treatment by an intelligent vacuum-assisted biopsy,” Eur. J. Cancer, vol. 143, pp. 134–146, Jan. 2021, doi: 10.1016/J.EJCA.2020.11.006.

[19] R. Haneef et al., “Use of artificial intelligence for public health surveillance: a case study to develop a machine Learning-algorithm to estimate the incidence of diabetes mellitus in France,” Arch. Public Heal., vol. 79, no. 1, pp. 1–13, 2021, doi: 10.1186/s13690-021-00687-0.

[20] M. Sharma, “Data Mining Prediction Techniques in Health Care Sector,” J. Phys. Conf. Ser., vol. 2267, no. 1, 2022, doi: 10.1088/1742-6596/2267/1/012157.

[21] “Python Logistic Regression Tutorial with Sklearn & Scikit | DataCamp.” https://www.datacamp.com/tutorial/understanding-logistic-regression-python (accessed Dec. 29, 2023).

[22] “Scikit-learn SVM Tutorial with Python (Support Vector Machines) | DataCamp.” https://www.datacamp.com/tutorial/svm-classification-scikit-learn-python (accessed Dec. 29, 2023).

[23] “Random Forest Classifier using Scikit-learn - GeeksforGeeks.” https://www.geeksforgeeks.org/random-forest-classifier-using-scikit-learn/ (accessed Dec. 29, 2023).

[24] “Gradient Boosting with Scikit-Learn, XGBoost, LightGBM, and CatBoost - MachineLearningMastery.com.” https://machinelearningmastery.com/gradient-boosting-with-scikit-learn-xgboost-lightgbm-and-catboost/ (accessed Dec. 29, 2023).

[25] “A Comprehensive Guide to Using Scikit-Learn’s MLPClassifier - AITechTrend.” https://aitechtrend.com/a-comprehensive-guide-to-using-scikit-learns-mlpclassifier/ (accessed Dec. 29, 2023).

[26] “Getting Started With The Extra Trees Algorithm In Python 2023.” https://hands-on.cloud/getting-started-with-the-extra-trees-algorithm-in-python/ (accessed Dec. 29, 2023).

[27] “Decision Tree Classifier with Sklearn in Python • datagy.” https://datagy.io/sklearn-decision-tree-classifier/ (accessed Dec. 29, 2023).

[28] “Bagging Classifier Python Code Example.” https://vitalflux.com/bagging-classifier-python-code-example/ (accessed Dec. 29, 2023).

[29] “K-nearest Neighbors in Scikit-learn - KDnuggets.” https://www.kdnuggets.com/2022/07/knearest-neighbors-scikitlearn.html (accessed Dec. 29, 2023).

[30] “Linear Discriminant Analysis Made Simple & How To Tutorial.” https://spotintelligence.com/2023/11/07/linear-discriminant-analysis-lda/ (accessed Dec. 29, 2023).

[31] P. Mohanty, L. Patnaik, G. Nayak, and A. Dutta, “Gender difference in prevalence of hypertension among Indians across various age-groups: a report from multiple nationally representative samples,” BMC Public Health, vol. 22, no. 1, pp. 1–10, 2022, doi: 10.1186/s12889-022-13949-5.

[32] P. Chhabra et al., “Gender-specific factors associated with hypertension among women of childbearing age: Findings from a nationwide survey in India,” Front. Cardiovasc. Med., vol. 9, no. December, pp. 1–9, 2022, doi: 10.3389/fcvm.2022.999567.

[33] T. J. Toney-Butler and J. M. Thayer, “Nursing Process,” Fundam. Nurs. Made Incred. Easy! Second Ed., p. 4, Apr. 2023, doi: 10.5005/jp/books/14252_4.

