## Supplementary material for "Applied machine learning techniques for chronic disease treatment default prediction and its potential benefits for patient outcome: A case series study approach": Request and approval letter

Kwame Nkrumah
University of Science
and Technology, Kumasi

College of Science

DEPARTMENT OF COMPUTER SCIENCE

Our Ref: DCS/S.1/Vol.1 Your Ref .................. Date: March 25, 2022

The Director
Kwahu Government Hospital
Atibie Kwahu
Eastern Region

KWAHU GOVERNMENT HOSPITAL RECEIVED DATE 30/03/22 ATIBIE

Dear Sir,

**COLLECTION OF DATA**

This is to introduce to you Mr. Michael Owusu-Adjei, PhD Computer Science student with index number PG20627433 of the Computer Science Department, Kwame Nkrumah University of Science and Technology.

He is undertaking a project work on the topic: "**Adherence Risk Prediction for Hypertensive Patients with Comorbidities Using Machine Learning Algorithms**" in partial fulfilment of the requirement for the award of his degree certificate and has chosen your outfit as his case study.

It would be greatly appreciated if you could give him the necessary assistance he may need.

The whole exercise will be of academic interest and any information provided will be treated as confidential.

Thank you for your co-operation and assistance.

for Prof. J. B. Hayfron-Acquah
**HEAD**

30/03/2022
Data collection approved.

DR. KOBENA A. WIREDU
(B.Sc MBChB. MGL)
MEDICAL SUPERINTENDENT
KWAHU GOV'T. HOSPITAL
MPRAESO - ATIBIE E/R.
